# Fasting hyperinsulinemia persists across type 2 diabetes duration without exogenous insulin

**DOI:** 10.64898/2026.07.27.26358997

**Authors:** Sana Mirza, Nadia Ernst, Joshua Moen

## Abstract

Declining insulin with longer type 2 diabetes (T2D) duration is commonly interpreted as beta-cell failure. We analyzed NHANES data from 2003–August 2023 cross-sectionally and 2007–2018 with mortality linkage. The primary harmonized analysis included 2,072 adults with diagnosed T2D, calculable duration, and no exogenous insulin, and 9,050 normoglycemic adults. Across durations of <5, 5–14, and ≥15 years, adjusted insulin ratios versus normoglycemia were 2.39 (95% CI 2.20–2.60), 1.88 (1.76–2.00), and 1.71 (1.58–1.86). A sensitivity restricted to current laboratory diabetes without exogenous insulin or secretagogues (n=808) yielded ratios of 2.21, 1.86, and 1.88. Among adults aged ≥75 years with HbA1c-confirmed T2D for ≥15 years and no exogenous insulin or secretagogue, the ratio was 1.73 (1.36–2.21; n=37). In the prespecified primary mortality analysis of the most recent six-cycle NDI-linked window, 2007–2018, insulin was associated with cardiovascular mortality in prediabetes (hazard ratio 2.95, 1.70–5.11). Hyperinsulinemia persisted across T2D duration and advanced age, arguing against uniform insulin deficiency in long-duration non-insulin-treated T2D. Mortality associations differed by disease stage and survey period.

## Introduction

Diabetes encompasses heterogeneous states of insulin availability and action, from absolute endogenous insulin deficiency to marked hyperinsulinemia accompanying severe insulin resistance. Hyperglycemia is the diagnostic phenotype shared by these states, but it does not reveal the physiology producing it. Similar glucose values can therefore arise from too little endogenous insulin, high insulin exposure, or both. This distinction matters for T2D and its precursors, which affect more than 130 million adults in the United States and impose substantial clinical and economic burdens [1,2].

Diagnosis and glycemic monitoring nevertheless remain organized around fasting glucose, oral glucose tolerance, and glycated hemoglobin (HbA1c) [3].

Fasting hyperinsulinemia often precedes glucose dysregulation and future T2D in prospective studies [4–6]. In established disease, however, declining insulin concentrations with longer reported duration are often interpreted as progressive beta-cell failure [7]. That interpretation is difficult to separate from aging, treatment selection, reduced hepatic insulin clearance, and survival to long disease duration. Serum insulin is not a direct measure of secretion, but it remains the hormone exposure experienced by insulin-responsive tissues. Whether the lower insulin observed with longer T2D duration reflects beta-cell failure or the changing composition of treated and surviving populations therefore remains unresolved.

We used nationally representative NHANES data to test whether fasting hyperinsulinemia persists across increasing durations of T2D in adults not using exogenous insulin. We hypothesized that fasting insulin would remain elevated relative to normoglycemia even after long disease duration and in advanced age. We also examined whether the association between fasting insulin and mortality differs between prediabetes and established T2D, where treatment, clearance, complications, and survival may alter what circulating insulin represents.

## Results

### Study populations

The cross-sectional source included 27,195 fasting-subsample adults across 11 nonoverlapping NHANES periods from 1999 through August 2023. Because no absolute conversion exists for the 1999–2002 radioimmunoassay, the primary harmonized analysis used 2003–August 2023. It included 9,050 normoglycemic adults and 2,072 adults with diagnosed T2D, calculable nonnegative duration, and no exogenous insulin; duration groups contained 695, 871, and 506 participants. An assay-robust sensitivity allowed 1999–2002 to contribute on its native log scale with survey-period fixed effects, increasing the T2D sample to 2,318. Drug-level prescription data ended in March 2020. The strict sensitivity within that window included 8,367 normoglycemic adults and 808 adults with current laboratory diabetes who used neither exogenous insulin nor an insulin secretagogue.

### Fasting insulin persists across diabetes duration

In the primary harmonized all-adult analysis, fasting insulin remained elevated at every T2D duration after exclusion of exogenous insulin (Table 1). Adjusted ratios versus normoglycemia were 2.39 (95% CI 2.20–2.60) before 5 years, 1.88 (1.76–2.00) at 5–14 years, and 1.71 (1.58–1.86) after at least 15 years. The strict sensitivity excluding secretagogues and requiring current laboratory diabetes produced ratios of 2.21 (1.99–2.45), 1.86 (1.69–2.04), and 1.88 (1.68–2.11), respectively. The assay-robust 1999–August 2023 sensitivity produced ratios of 2.27, 1.79, and 1.61. Thus, the lower insulin concentration observed with duration in the full sample did not approach normoglycemia and was absent after restriction to biochemically active diabetes without pharmacologic stimulation of secretion.

**Table 1.** Fasting insulin across T2D duration in the primary harmonized cohort and strict sensitivity analysis, NHANES 2003–August 2023.

| Group | Primary: no<br>exogenous<br>insulin, n | Adjusted ratio (95%<br>CI) | Strict<br>sensitivity,<br>n | Adjusted ratio (95%<br>CI) |
| --- | --- | --- | --- | --- |
| Normoglycemia | 9,050 | 1.00 (reference) | 8,367 | 1.00 (reference) |
| T2D <5 years | 695 | 2.39 (2.20–2.60) | 284 | 2.21 (1.99–2.45) |
| T2D 5–14 years | 871 | 1.88 (1.76–2.00) | 346 | 1.86 (1.69–2.04) |
| T2D ≥15 years | 506 | 1.71 (1.58–1.86) | 178 | 1.88 (1.68–2.11) |

The primary harmonized analysis required diagnosed T2D with calculable nonnegative duration and excluded exogenous insulin; oral therapies, including secretagogues, remained eligible. The strict 2003–March 2020 sensitivity additionally excluded sulfonylureas, meglitinides, dipeptidyl-peptidase-4 inhibitors, and glucagon-like peptide-1 receptor agonists and required HbA1c ≥6.5% or fasting glucose ≥126 mg/dL. Ratios are survey-weighted and adjusted for age, sex, race/ethnicity, and survey period. All comparisons had $P < 1 \times 10^{-1}$ . CI, confidence interval; T2D, type 2 diabetes.

### Aging, frailty, and the oldest long-duration group

The primary harmonized pattern remained present within the older population. Among adults aged 60 years or older, adjusted ratios versus normoglycemia were 2.05 before 5 years, 1.84 at 5–14 years, and 1.63 after at least 15 years. Fasting insulin declined with age in normoglycemic adults as well as in T2D (Table 2). The normoglycemic slope was −0.51% per year. The 5–14-year T2D slope was −0.38% per year and did not differ from normoglycemia (P=0.60). Slopes were steeper for T2D of less than 5 years (−1.32% per year; P=0.002 versus normoglycemia) and at least 15 years (−1.40% per year; P=0.026), although the latter slope rests on six participants aged 18–39, reported in Table 2 as uninterpretable.

**Table 2.** Age-related fasting-insulin concentrations and slopes, NHANES 2003–August 2023.

| Group | 18–39 years | 40–59 years | ≥60 years | Slope,<br>%/year | P vs<br>normoglycemia |
| --- | --- | --- | --- | --- | --- |
| Normoglycemia | 7.00<br>(n=5,041) | 5.91<br>(n=2,474) | 5.81<br>(n=1,535) | −0.51 | Reference |
| T2D <5 years | 21.84 (n=61) | 15.31<br>(n=290) | 12.17<br>(n=344) | −1.32 | 0.002 |
| T2D 5–14 years | 12.15 (n=39) | 11.53<br>(n=292) | 10.98<br>(n=540) | −0.38 | 0.60 |
| T2D ≥15 years | Not<br>interpreted<br>(n=6) | 12.09 (n=78) | 9.82 (n=422) | −1.40 | 0.026 |

Cells are survey-weighted geometric means (μU/mL); slopes are from ln(fasting insulin) modeled continuously against age, adjusted for sex, race/ethnicity, and survey period. Participants using exogenous insulin were excluded from T2D groups.

Adjustment for body mass index and a frailty-proxy set comprising low body mass, hypoalbuminemia, inactivity, prevalent cardiovascular disease or cancer, and estimated glomerular filtration rate below 60 mL/min/1.73 m^2^ attenuated but did not remove the ≥15-year association in adults aged 60 years or older (ratio 1.33, 95% CI 1.22–1.44; Supplementary Table S2). In the separate physical-function sample, adding walking, stair-climbing, chair-rise, and mobility-equipment limitations produced a ratio of 1.31 (1.19–1.44); restricting to participants reporting no functional difficulty produced 1.33 (1.16– 1.52).

The oldest and most restricted subgroup showed the same result. Among adults aged 75 years or older with HbA1c ≥6.5%, at least 15 years since diagnosis, no exogenous insulin, and no secretagogue, geometric-mean fasting insulin was 10.92 versus 5.58 µU/mL in normoglycemic comparators; the adjusted ratio was 1.73 (95% CI 1.36–2.21; P=4.7×10LJLJ; n=37).

### Treatment context

Across adults aged 60 years or older in the 2003–August 2023 cross-sectional cohort, geometric-mean fasting insulin was 5.81 µU/mL in normoglycemia, 9.21 µU/mL in prediabetes, 11.93 µU/mL in non-insulin-treated T2D, and 17.12 µU/mL in insulin-treated T2D (Supplementary Table S1).

### Fasting insulin and cardiovascular mortality

For the secondary mortality aim, the primary analysis included 4,021 adults with prediabetes (weighted population: 68.5 million) and 1,385 with oral-therapy T2D (weighted population: 16.8 million) in the most recent six-cycle NDI-linked window, NHANES 2007–2018 (Table 3). In prediabetes, each one-natural-log-unit higher fasting insulin was associated with cardiovascular mortality (design-robust HR 2.95, 95% CI 1.70–5.11; P=1.2×10□□) and all-cause mortality (HR 1.69, 1.15–2.47). Cancer mortality was null in this window. Historical and pooled cohorts were sensitivity analyses. The prediabetes cardiovascular association was not reproduced in 1999–2006 (HR 0.80, 0.42–1.53; P=0.50), and the pooled 1999–2018 estimate was not significant (HR 1.46, 0.90–2.37; P=0.124). The independent between-period contrast was significant (P=0.0026). In oral-therapy T2D, the primary 2007–2018 cardiovascular estimate was borderline and inverse (HR 0.51, 0.26–1.00; P=0.049), but it was not significant under competing-risk analysis or when 1999–2018 cycles were pooled (HR 0.84, 0.54– 1.31; P=0.45; Supplementary Table S3).

**Table 3.**
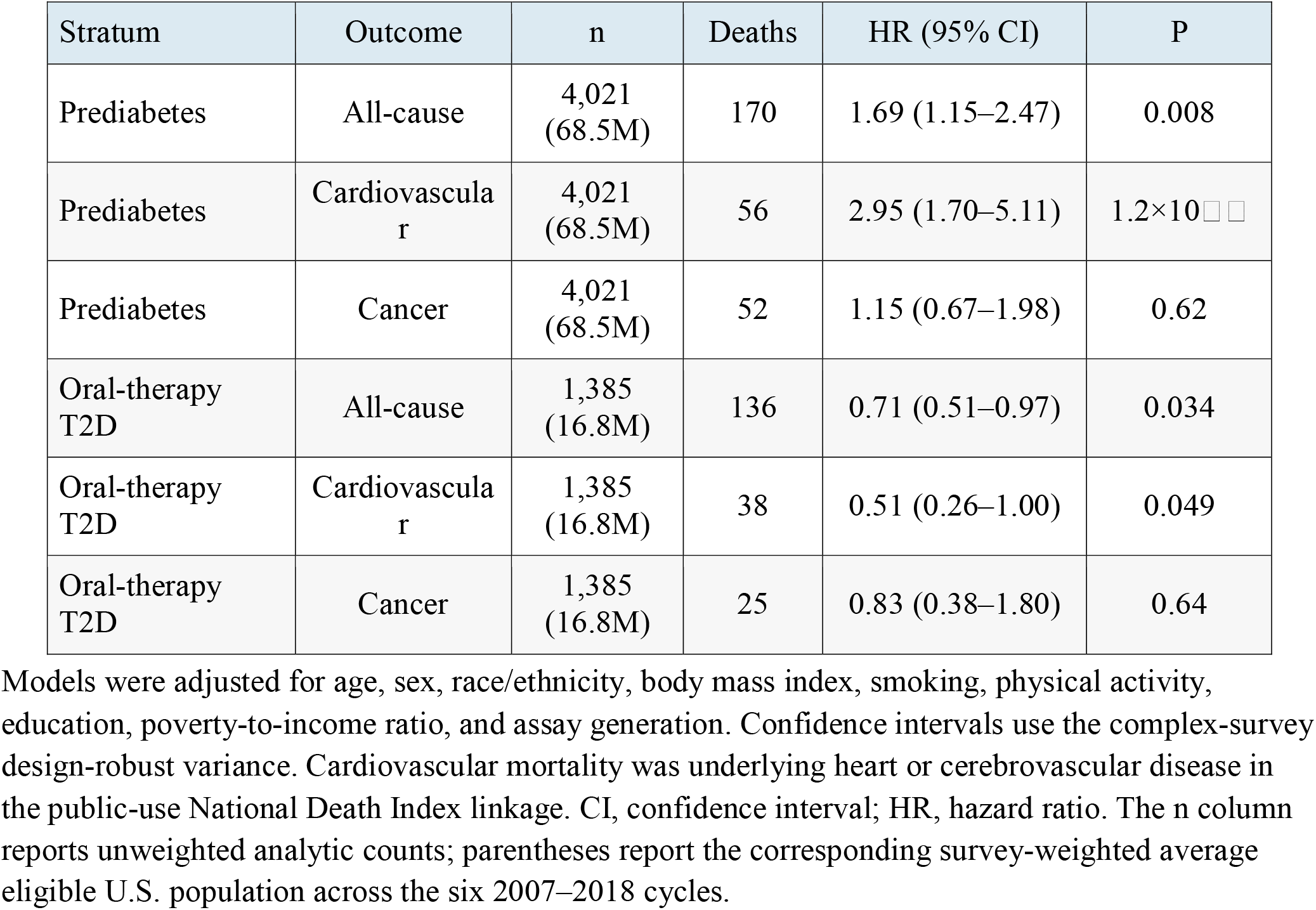
Survey-weighted mortality associations per one-natural-log-unit higher fasting insulin, NHANES 2007–2018.

| Stratum | Outcome | n | Deaths | HR (95% CI) | P |
| --- | --- | --- | --- | --- | --- |
| Prediabetes | All-cause | 4,021<br>(68.5M) | 170 | 1.69 (1.15–2.47) | 0.008 |
| Prediabetes | Cardiovascular | 4,021<br>(68.5M) | 56 | 2.95 (1.70–5.11) | 1.2×10<sup>−4</sup> |
| Prediabetes | Cancer | 4,021<br>(68.5M) | 52 | 1.15 (0.67–1.98) | 0.62 |
| Oral-therapy<br>T2D | All-cause | 1,385<br>(16.8M) | 136 | 0.71 (0.51–0.97) | 0.034 |
| Oral-therapy<br>T2D | Cardiovascular | 1,385<br>(16.8M) | 38 | 0.51 (0.26–1.00) | 0.049 |
| Oral-therapy<br>T2D | Cancer | 1,385<br>(16.8M) | 25 | 0.83 (0.38–1.80) | 0.64 |

Models were adjusted for age, sex, race/ethnicity, body mass index, smoking, physical activity, education, poverty-to-income ratio, and assay generation. Confidence intervals use the complex-survey design-robust variance. Cardiovascular mortality was underlying heart or cerebrovascular disease in the public-use National Death Index linkage. CI, confidence interval; HR, hazard ratio. The n column reports unweighted analytic counts; parentheses report the corresponding survey-weighted average eligible U.S. population across the six 2007–2018 cycles.

## Discussion

The primary finding is that harmonized fasting insulin persisted across T2D duration in adults not using exogenous insulin. Adjusted ratios versus normoglycemia were 2.39 before 5 years, 1.88 at 5–14 years, and 1.71 after at least 15 years. Secretagogues remained eligible in this analysis because circulating insulin still reflected endogenous pancreatic output. In the strict sensitivity excluding secretagogues and requiring current laboratory diabetes, ratios were 2.21, 1.86, and 1.88. The full-cohort decline with duration therefore did not approach normoglycemia, and the decline disappeared when the analysis was restricted to ongoing biochemical diabetes without pharmacologic stimulation of secretion. The assay-robust model including 1999–2002 produced the same interpretation.

The secondary aging analyses show that the full-cohort finding is not explained by the age distribution. Harmonized ratios remained 2.05, 1.84, and 1.63 among adults aged 60 years or older. Fasting insulin fell with age in normoglycemic adults, and the 5–14-year T2D slope was similar to that background decline (P=0.60). The association persisted after proxy adjustment for frailty and functional limitation and remained present in adults aged at least 75 years. Populations reaching long-duration T2D without exogenous insulin therefore remain hyperinsulinemic relative to their normoglycemic contemporaries.

These findings clarify why glucose alone may be insufficient to characterize the physiology relevant to glycemic intensification in older adults. Participants receiving exogenous insulin had the highest measured fasting insulin concentration in this analysis, 17.12 versus 11.93 µU/mL in non-insulin-treated T2D, although the assay could not distinguish exogenous from endogenous insulin in users.

Glucose identifies failed glycemic homeostasis without determining whether endogenous insulin is deficient, preserved, or elevated. A physiology-based framework would distinguish insulin replacement for demonstrated deficiency from strategies that reduce insulin demand while maintaining safe glycemia when endogenous insulin is preserved or elevated. These data do not establish fasting-insulin thresholds for selecting therapy, but they support studying insulin state as a dimension of diabetes heterogeneity rather than treating a hyperinsulinemic state with increased insulin.

In the primary NDI-linked 2007–2018 mortality analysis, each one-natural-log-unit higher fasting insulin was associated with cardiovascular mortality within prediabetes (HR 2.95, 95% CI 1.70–5.11). Experimental and clinical studies link hyperinsulinemia to vascular, renal, and sympathetic pathways [8–13].

The contrast between prediabetes and established T2D may reflect a stage-dependent change in what fasting insulin represents. In prediabetes, endogenous hyperinsulinemia may more directly index insulin resistance and vascular pathways that precede overt disease. After T2D is established, circulating insulin increasingly reflects treatment selection, residual beta-cell function, renal and hepatic clearance, accumulated complications, and survival. The borderline inverse estimate in oral-therapy T2D disappeared in pooled and competing-risk analyses and should not be interpreted as protective. Instead, it is compatible with progressive separation of measured insulin from the causal components represented by hyperinsulinemia earlier in the disease course.

Strengths include a nationally representative primary cohort spanning the adult age range with harmonized insulin from 2003 through August 2023, an assay-robust sensitivity extending to 1999, exclusion of exogenous insulin, and a stringent laboratory-active sensitivity excluding secretagogues. Age-restricted, frailty, physical-function, kidney-function, and advanced-age analyses further tested persistence. The duration comparison is cross-sectional and therefore describes populations rather than individual trajectories. Further limitations include self-reported age at diagnosis, a single fasting measurement, absence of clamp-derived insulin sensitivity, and incomplete drug-level medication data after March 2020. Fasting insulin reflects both secretion and clearance, and medication use and survival to long disease duration may select different disease phenotypes. The drug-restricted group aged 75 years or older was small. For the mortality analysis, the association varied across assay eras and harmonization choices, the oral-therapy upper confidence limit reached 1.00, and the pooled 1999– 2018 estimate was null.

Across the adult population, long-duration T2D without exogenous insulin remained hyperinsulinemic relative to age-adjusted normoglycemia. The finding persisted after excluding secretagogues, requiring current laboratory diabetes, and restricting analyses to older and frailty-adjusted populations. The differing mortality associations across prediabetes and established T2D support further study of how treatment, clearance, complications, and survival alter the meaning of circulating insulin across disease stages. These observational data do not establish a treatment target or support routine fasting-insulin testing.

## Methods

### Study design and data windows

NHANES is a continuous, multistage probability sample of the non-institutionalized United States civilian population. We combined 11 non-overlapping periods: 1999–2000, 2001–2002, eight subsequent releases through the 2017–March 2020 pre-pandemic file, and August 2021–August 2023. Non-pregnant adults aged 18 years or older were eligible if they completed the mobile examination center fasting protocol, had a positive fasting-subsample weight, and had a measured fasting insulin concentration. The primary harmonized duration analysis used 2003–August 2023. An assay-robust sensitivity added 1999–2002 on its native log scale, and the medication-class sensitivity ended in March 2020 because public drug-level prescription data were unavailable for 2021–2023.

### Insulin measurement and harmonization

The primary laboratory exposure was fasting serum or plasma insulin. The 1999–2002 double-antibody radioimmunoassay has no defensible absolute bridge to later platforms and was excluded from absolute-concentration analyses, although it was retained in cycle-fixed-effect sensitivity models on native log-insulin. Values from 2005–2010 Mercodia assays were converted to the Tosoh scale as 1.0526×Mercodia−1.5674. Roche 2011–2012 values were converted as 10^(1.024×log10[Roche]−0.0802). The 2003–2004 and 2013 onward files used Tosoh-platform values. The 2009–2010 collection changed assay during fieldwork; the cycle conversion is therefore an approximation and survey-period fixed effects were retained in regression models.

### Diabetes duration and primary comparison

Self-reported T2D required a physician diagnosis and age at diagnosis. Duration was current age minus reported age at diagnosis and was grouped as less than 5, 5–14, or at least 15 years. Normoglycemia required no self-reported diabetes, HbA1c <5.7%, and fasting glucose <100 mg/dL. The primary analysis included all adults aged at least 18 years with calculable T2D duration and no reported or prescribed exogenous insulin. Oral glucose-lowering therapies, including secretagogues, remained eligible because measured circulating insulin was endogenous. The strict sensitivity required current laboratory diabetes, defined as HbA1c ≥6.5% or fasting glucose ≥126 mg/dL, and excluded both exogenous insulin and prescribed drugs that directly stimulate insulin secretion. Secretagogues comprised sulfonylureas, meglitinides, dipeptidyl-peptidase-4 inhibitors, and glucagon-like peptide-1 receptor agonists. Drug classes were derived from NHANES prescription identifiers linked to the public drug lexicon. Metformin, sodium-glucose cotransporter-2 inhibitors, thiazolidinediones, and alpha-glucosidase inhibitors were not classified as secretagogues.

### Aging, frailty, and function analyses

Age groups were 18–39, 40–59, and at least 60 years. Aging slopes were estimated by interaction between continuous age and duration group after exclusion of exogenous-insulin users. Frailty proxies were body mass index below 20 kg/m^2^, albumin below 3.5 g/dL, inactivity, prevalent cardiovascular disease or cancer, and estimated glomerular filtration rate below 60 mL/min/1.73 m^2^, calculated with the 2021 CKD-EPI race-free equation [14]. A separate 1999–2018 physical-function analysis used reported difficulty walking one-quarter mile, climbing ten steps, rising from an armless chair, and mobility-equipment use. The strict oldest subgroup required age at least 75 years, HbA1c ≥6.5%, duration at least 15 years, and no exogenous insulin or secretagogue.

### Mortality cohort

For the secondary mortality aim, the primary analysis used the most recent six complete pre-pandemic NHANES cycles with public-use National Death Index linkage: 2007–2018 recruitment linked through 31 December 2019. This prespecified contemporary window bounded follow-up to approximately a decade and limited the distance from modern diagnostic and treatment practice. A historical 1999– 2006 analysis and a pooled 1999–2018 analysis were retained as temporal-robustness sensitivities rather than primary cohorts. Because the 1999–2002 radioimmunoassay has no absolute bridge to later platforms, all-cycle estimates per one natural-log unit require caution; a separate 2003–2018 analysis standardized log insulin within survey period. Participants with prevalent cardiovascular disease or cancer were excluded. Prediabetes required HbA1c 5.7–6.4% or fasting glucose 100–125 mg/dL without glucose-lowering therapy. Oral-therapy T2D excluded exogenous insulin and included self-reported or laboratory diabetes or non-insulin glucose-lowering therapy. Cardiovascular mortality was underlying heart disease or cerebrovascular disease; cancer mortality was a secondary comparator.

### Statistical analysis

All estimates incorporated NHANES strata, pseudo-primary sampling units, and pooled fasting-subsample weights scaled to the number of survey years contributed by each period. Analyses used complete cases for variables required by each model; model-specific sample sizes are reported in the tables. Geometric means were exponentiated survey-weighted means of log insulin. Primary duration ratios were obtained from survey-weighted linear regression of harmonized 2003–August 2023 log insulin with adjustment for age, sex, race/ethnicity, and survey period. The model was repeated by age group. An assay-robust sensitivity used native log insulin with period fixed effects, allowing 1999– 2002 to contribute without absolute conversion. The strict laboratory-active, no-secretagogue sensitivity used harmonized 2003–March 2020 insulin values with the same covariate adjustment.

Age-slope models included age-by-duration-group interactions. Additional sensitivity models added body mass index, frailty proxies, or functional burden as specified. Mortality models used survey-weighted Cox proportional hazards with design-robust variance and adjustment for age, sex, race/ethnicity, body mass index, smoking, physical activity, education, poverty-to-income ratio, and assay generation. Hazard ratios are per one-natural-log-unit higher fasting insulin. Survey-weighted Fine–Gray models treated non-cardiovascular death as a competing event. Proportionality was assessed with scaled Schoenfeld residuals from unweighted Cox models fit to the aligned analytic sample; no violation was detected for fasting insulin (all exposure P≥0.183; all global P≥0.176). Mortality outcomes were treated as separate secondary comparisons, and P values were not adjusted for multiple testing. All tests were two-sided with a nominal alpha level of 0.05. Analyses were performed in R and Stata MP 18.0.

### Ethics and use of artificial intelligence

NHANES protocols were approved by the National Center for Health Statistics Research Ethics Review Board, and all participants provided written informed consent. This secondary analysis used de-identified public data and required no additional institutional review. Data were accessed from June 2025 through June 2026 without exposure to information that could identify participants. The study was conducted in accordance with applicable regulations and the Declaration of Helsinki. Generative artificial intelligence (OpenAI) was used for formatting assistance and code review; the authors independently reviewed and verified all analyses, numerical results, interpretations, and final text. No artificial-intelligence tool is an author.

## Data Availability

This analysis used de-identified, publicly available data from the National Health and Nutrition Examination Survey (NHANES), conducted by the National Center for Health Statistics (NCHS).

https://wwwn.cdc.gov/nchs/nhanes/

## Data availability

NHANES examination, laboratory, questionnaire, prescription, and public-use linked mortality files are available from the National Center for Health Statistics at https://www.cdc.gov/nchs/nhanes/ and https://www.cdc.gov/nchs/data-linkage/mortality-public.htm. No new participant-level dataset was generated for this study.

## Code availability

The versioned R analysis code is available upon request. The archive contains no restricted or participant-level data.

## Acknowledgements

The authors thank the NHANES participants and the National Center for Health Statistics staff who collected and released the data.

## Author contributions

J.M. conceived the study, designed and performed the analysis, interpreted the results, and supervised the work. S.M. contributed to data curation, interpretation, and manuscript drafting. N.E. contributed to data curation, interpretation, and critical revision. All authors reviewed and approved the manuscript.

## Additional Information

