## Supplemental Information for "Fasting hyperinsulinemia persists across type 2 diabetes duration without exogenous insulin"

### Supplementary Table S1. Fasting insulin across glycemic and treatment stages among adults aged 60 years or older, NHANES 2003–August 2023.

| Glycemic/treatment group | n | Geometric mean, µU/mL (95% CI) |
| --- | --- | --- |
| Normoglycemia | 1,535 | 5.81 (5.56–6.07) |
| Prediabetes | 3,816 | 9.21 (8.93–9.49) |
| Non-insulin-treated T2D | 2,050 | 11.93 (11.49–12.39) |
| Insulin-treated T2D | 483 | 17.12 (14.98–19.57) |

Values are survey-weighted. Normoglycemia required no self-reported diabetes, HbA1c <5.7%, and fasting glucose <100 mg/dL. Prediabetes required HbA1c 5.7–6.4% or fasting glucose 100–125 mg/dL without glucose-lowering therapy. T2D included self-reported or laboratory diabetes or glucose-lowering therapy.

### Supplementary Table S2. Frailty-proxy and physical-function sensitivities among adults aged 60 years or older.

| Model | <5 years | 5–14 years | ≥15 years |
| --- | --- | --- | --- |
| Frailty-complete (n=3,367); age/sex/race/period adjusted | 2.06 (1.88–2.25) | 1.80 (1.67–1.93) | 1.62 (1.50–1.75) |
| Additionally adjusted for BMI and frailty proxies (n=3,367) | 1.63 (1.50–1.76) | 1.44 (1.36–1.53) | 1.33 (1.22–1.44) |
| Zero frailty-proxy deficits; additionally adjusted for BMI (n=859) | 1.59 (1.33–1.91) | 1.46 (1.23–1.73) | 1.32 (1.11–1.58) |
| Physical-function complete; additionally adjusted for BMI and burden (n=2,192) | 1.48 (1.34–1.64) | 1.39 (1.29–1.49) | 1.31 (1.19–1.44) |
| No reported functional difficulty; additionally adjusted for BMI (n=1,206) | 1.45 (1.26–1.65) | 1.39 (1.25–1.55) | 1.33 (1.16–1.52) |

Entries are adjusted fasting-insulin ratios versus age-adjusted normoglycemic adults aged 60 years or older. Frailty proxies were BMI <20 kg/m², albumin <3.5 g/dL, inactivity, prevalent cardiovascular disease or cancer, and eGFR <60 mL/min/1.73 m². Functional burden included difficulty walking one-quarter mile, climbing ten steps, rising from an armless chair, and mobility-equipment use.

### Supplementary Table S3. Cardiovascular-mortality window and competing-risk sensitivities.

| Stratum | Window/model | Events | Exposure scale | Estimate (95% CI) | P |
| --- | --- | --- | --- | --- | --- |
| Prediabetes | 2007–2018 Cox n=4,021 (68.5M weighted) | 56 | +1 ln-unit | 2.95 (1.70–5.11) | 1.2×10⁻⁴ |
| Prediabetes | 1999–2018 Cox (n=5,641) | 139 | +1 ln-unit | 1.46 (0.90–2.37) | 0.124 |
| Prediabetes | 2003–2018 assay-robust Cox (n=4,870) | 94 | +1 within-cycle SD | 1.68 (1.23–2.30) | 0.0012 |
| Oral-therapy T2D | 2007–2018 Cox n=1,385 (16.8M weighted) | 38 | +1 ln-unit | 0.51 (0.26–1.00) | 0.049 |
| Oral-therapy T2D | 1999–2018 Cox (n=1,931) | 110 | +1 ln-unit | 0.84 (0.54–1.31) | 0.45 |
| Oral-therapy T2D | 2007–2018 Fine–Gray n=1,385 (16.8M weighted) | 38 | +1 ln-unit | 0.58 (0.29–1.17) | 0.127 |

The 1999–2002 assay is not directly convertible to the later Tosoh scale. The 2003–2018 assay-robust model standardized log insulin within survey period; its scale therefore differs from the +1 ln-unit models. The independent prediabetes contrast between 1999–2006 and 2007–2018 yielded P=0.0026. Fine–Gray analysis treated noncardiovascular death as a competing event. Confidence intervals use design-robust variance. For 2007–2018 rows, the value in parentheses after n is the survey-weighted average eligible U.S. population across the six survey cycles.
